# Normative Speech Modeling for ALS Diagnosis with Application to Other Neurodegenerative Diseases

**DOI:** 10.64898/2026.05.25.26354057

**Authors:** Mithil Shah

## Abstract

Amyotrophic lateral sclerosis (ALS) is a progressive neurodegenerative disease affecting more than 450,000 individuals worldwide and is frequently diagnosed more than 12 months after symptom onset, delaying intervention during a critical early window. Because up to 80% of patients develop dysarthria within two years, subtle changes in speech provide a signal of early bulbar motor neuron degeneration. However, existing speech-based systems rely on supervised classification trained on limited datasets, achieving moderate sensitivity and depending heavily on labeled disease examples, which restrict scalability and early detection. This study introduces SPEAK-NORM, the first-ever normative speech modeling framework for early ALS diagnosis, which learns age- and sex-conditioned motor-speech distributions exclusively from healthy individuals. A conditional variational autoencoder models coordination of hypoglossal, laryngeal, and respiratory motor pathways, and deviation from this healthy manifold is quantified through latent representations and reconstruction error to form a 354-dimensional profile. A calibrated linear Support Vector Machine performs subject-level classification under subject-disjoint validation. On the VOC-ALS database (n = 153), SPEAK-NORM achieves 98% accuracy with balanced sensitivity and specificity, significantly outperforming established clinical acoustic indices and prior systems. The framework maintains strong performance under cross-task generalization and when retrained on healthy controls in independent dementia and Parkinson’s disease cohorts, demonstrating disease-specific deviation patterns rather than generic neurodegenerative change. Spectral, temporal, and latent separations further support interpretability. By modeling healthy speech instead of memorizing disease examples, SPEAK-NORM enables scalable early neuromotor screening using recording devices, with potential to support earlier diagnosis, differential classification, and monitoring of ALS progression.

## 1 Purpose and Hypothesis

This study investigates whether age- and sex-conditioned normative generative modeling of healthy speech can enable earlier and more precise detection of ALS-related motor speech degeneration, distinguish ALS-specific deviation from other neurodegenerative disorders with overlapping speech abnormalities, and stratify ALS patients into clinically meaningful motor speech phenotypes aligned with ALSFRS-R speech scores. The central hypothesis is that if a conditional generative model is trained exclusively on healthy speech to learn the structured distribution of normative motor coordination, then individuals with ALS will exhibit statistically significant spectral, temporal, and latent deviations from this healthy manifold that improve diagnostic sensitivity beyond fixed acoustic thresholds, demonstrate disorder-specific deviation patterns relative to Parkinson’s disease and dementia, and reveal distinct bulbar motor-speech subtypes through clustering of subject-level deviation profiles.

## 2 Background Research

Accurate and differential diagnosis of amyotrophic lateral sclerosis (ALS) remains a central challenge in clinical neurology (Paganoni et al., 2014). ALS affects more than 450,000 individuals worldwide and carries a median survival of two to four years after symptom onset (Young Faces of ALS, 2026). Diagnosis requires demonstration of progressive upper and lower motor neuron involvement across anatomical regions (Pugdahl et al., 2021). The Gold Coast Criteria increase sensitivity to 88.2%; however, confirmation depends on electromyographic abnormalities that may emerge months apart across spinal or bulbar segments (Makki and Benatar, 2007; Pugdahl et al., 2021). During this interval, neurodegeneration continues (Mejzini et al., 2019).

Importantly, early bulbar degeneration may produce subtle but structured acoustic alterations prior to exceeding fixed clinical thresholds or producing perceptible dysarthria (Milella et al., 2023). Threshold-based metrics such as jitter, shimmer, and diadochokinetic rate confirm abnormality only after scalar values surpass predefined cutoffs (Kent et al., 1999), potentially delaying recognition of evolving motor unit loss. A modeling framework capable of quantifying structured deviation from healthy motor-speech distributions may therefore enable detection of pre-threshold abnormalities, supporting earlier identification of bulbar involvement.

Furthermore, Parkinson’s disease and dementia produce overlapping bulbar and motor features, complicating differential classification (Aslam et al., 2023; Ferrari et al., 2011). Methods that distinguish ALS-specific motor pathway degeneration from generalized neuromotor impairment are therefore required to improve diagnostic specificity.

Bulbar motor neuron degeneration provides a defined anatomical substrate for addressing this requirement (Hoerter and Chandran, 2020). Approximately 25% of patients present with speech impairment at onset, and up to 75% develop dysarthria within two years (Target ALS, 2025; Young Faces of ALS, 2026). Lower motor neurons in the hypoglossal nucleus innervate the genioglossus and intrinsic lingual muscles, which coordinate articulatory precision (Hoerter and Chandran, 2020). Their denervation reduces movement velocity and increases articulatory timing variability (Pawlukowska et al., 2019). Motor neurons in the nucleus ambiguus innervate intrinsic laryngeal muscles that regulate vocal fold tension and adduction (Hoerter and Chandran, 2020); their degeneration increases jitter and shimmer and reduces pitch variability (Milella et al., 2023). Spinal motor neurons supplying the diaphragm and intercostals maintain subglottal pressure, and their loss reduces vocal intensity and phrase duration (Pawlukowska et al., 2019). Acoustic analysis has been shown to detect these perturbations prior to consistent perceptual recognition (Milella et al., 2023). Bulbar degeneration therefore produces a structured acoustic phenotype corresponding to specific motor pathway loss rather than nonspecific vocal weakness.

Beyond early detection, the heterogeneity of ALS motor involvement necessitates methods capable of stratifying patients by dominant motor-speech phenotype rather than reducing impairment to a single scalar severity score (de Carvalho and Swash, 2024). Because hypoglossal, laryngeal, and respiratory motor pathways contribute distinct acoustic consequences when degenerating (Hoerter and Chandran, 2020; Pawlukowska et al., 2019), deviation modeling may allow identification of pathway-specific patterns, supporting clinically meaningful stratification of bulbar phenotypes and progression trajectories.

Current clinical voice assessment in ALS relies on the KayPENTAX Multidimensional Voice Program (MDVP), which extracts jitter, shimmer, and noise-to-harmonic ratio (NHR) from sustained phonation, and diadochokinetic rate assessment, which compares syllable repetition speed against population norms (Kent et al., 1999). Both tools reduce the acoustic output of the vocal tract to a multidimensional parameter set compared against fixed normative thresholds. They do not account for the fact that the hypoglossal nucleus, nucleus ambiguus, and spinal respiratory motor neurons each produce distinct acoustic consequences when they degenerate (Hoerter and Chandran, 2020; Pawlukowska et al., 2019). When jitter rises above 1.04%, the measurement confirms laryngeal instability but does not reveal which nucleus lost motor units or how far denervation has progressed. A patient whose hypoglossal nucleus is actively degenerating but whose jitter has not yet crossed threshold will be assessed as normal, and lower motor neurons will continue to die (Paganoni et al., 2014; Mejzini et al., 2019). Research has consistently shown that healthy vocal parameters vary substantially with age and sex, yet the MDVP’s built-in normative thresholds are not stratified by these variables (Kent et al., 1999), meaning that deviations in elderly speakers may be misclassified as pathological while early perturbation changes in younger patients may fall within a falsely broad normal range. Further, Parkinson’s disease and frontotemporal dementia cross the same thresholds through entirely distinct mechanisms (Aslam et al., 2023; Ferrari et al., 2011), thereby raising the need to evaluate motor speech as a structured profile across the anatomically distinct pathways that ALS preferentially destroys, rather than as a single degradation index (de Carvalho and Swash, 2024).

Understanding the acoustic contribution of each bulbar motor pathway to healthy speech may be essential to achieving early and specific ALS diagnosis. The hypoglossal nucleus, nucleus ambiguus, and spinal respiratory motor neurons fire in coordinated sequence during connected speech, and that coordination produces a structured acoustic output reflecting the integrity of each population independently (Hoerter and Chandran, 2020; Pawlukowska et al., 2019). When one population begins to denervate, the resulting departure from healthy output is not a uniform reduction in vocal quality but a directional change across a specific subset of acoustic dimensions corresponding to that nucleus (Milella et al., 2023). Evaluating such changes therefore requires a reference model of healthy motor speech for a given speaker’s age and sex, against which patient recordings can be compared across all dimensions simultaneously. Normative modeling frameworks in neuroimaging have demonstrated that constructing reference distributions from healthy individuals and quantifying subject-level departure from them reveals pathway-specific involvement that scalar biomarkers cannot resolve (Bozek et al., 2023). To learn the high-dimensional structure of healthy motor speech required for such a reference, deep generative models can be leveraged for their ability to estimate full probability distributions rather than isolated acoustic indices (Woodland et al., 2025). Conditional variational autoencoders are particularly suited to this requirement because they learn age- and sex-conditioned distributions of healthy speech, enabling each recording to be evaluated as a structured departure from the normative output expected for that specific speaker (Ramchandran et al., 2023).

Conditional variational autoencoders (cVAEs) are a class of deep generative neural networks that learn structured latent distributions conditioned on observed covariates (Ramchandran et al., 2023). Unlike discriminative classifiers that separate labeled groups, cVAEs model the underlying distribution of healthy data, thereby capturing the full structure of normative biological variation. For example, normative modeling frameworks in neuroimaging employ related generative architectures to characterize subject-level deviation from population reference distributions across disorders (Bozek et al., 2023). For motor speech analysis, a cVAE may offer improved characterization of pathway-specific degeneration if designed to learn the conditioned distribution of healthy acoustic coordination across bulbar motor systems. By treating each speech recording as a sample drawn from an age- and sex-conditioned distribution, the cVAE evaluates its representation in the context of the healthy manifold, quantifying structured departure rather than scalar degradation.

In this study, a generative framework, SPEAK-NORM—**S**peech **P**athology **E**valuation through **A**utomated **K**nowledge of **NORM**ative Patterns—is trained exclusively on healthy recordings from the VOC-ALS database, a clinically validated and demographically diverse acoustic corpus (Bionetworks, 2026). This normative approach allows the model to learn the distribution of healthy motor speech across speakers without exposure to pathological data. SPEAK-NORM adopts a conditional variational autoencoder (cVAE) architecture because of its ability to model latent distributions conditioned on age and sex. In motor speech analysis, the cVAE evaluates each recording in the context of the conditioned healthy manifold, enabling structured characterization of deviation from normative output.

The VOC-ALS database comprises 153 subjects and 1,294 recordings spanning sustained phonation and diadochokinetic tasks, evaluated under subject-disjoint five-fold cross-validation (Bionetworks, 2026). The resultant deviation representations from SPEAK-NORM were compared against five established clinical acoustic baselines and three published state-of-the-art systems. Clinical comparators included eGeMAPS (Eyben et al., 2016), Dysphonia Severity Index (DSI) (Wuyts et al., 2000), F2 slope analysis (Yunusova et al., 2012), Envelope Modulation Spectrum (EMS) features (Liss et al., 2010), and the combined Formant Centralization Ratio with Bark-scaled Vowel Space Area (FCR + VSA-Bark) (Sapir et al., 2010). DSI, F2 slope, and FCR + VSA-Bark were evaluated on sustained vowel tasks only, whereas eGeMAPS and EMS were evaluated across all tasks in accordance with their methodological design (Eyben et al., 2016; Liss et al., 2010). In addition, performance was compared against three published state-of-the-art pathological speech detection systems: the SVM-based VOC-ALS classifier of Tena et al. (Tena et al., 2023), the hypernetwork architecture of Ilias et al. (Ilias and Askounis, 2025), and the diadochokinetic-based ALS detection framework of Agurto et al. (Agurto et al., 2024). Performance was assessed using ROC-AUC, PR-AUC, balanced accuracy, sensitivity, specificity, accuracy, and F1 score under identical subject-disjoint conditions. Cross-disorder generalization was further evaluated on independent dementia (n = 145; 455 recordings; 84 dementia; 61 no-dementia) (Gite, 2022) and Parkinson’s disease (n = 65) (Dimauro, 2019) cohorts to determine whether learned representations reflect ALS-specific motor pathway loss rather than generalized neurodegeneration. These evaluations position normative deviation modeling, as implemented in SPEAK-NORM, as a biologically grounded framework for accurate and differential ALS diagnosis.

## 3 Materials and Methods

### 3.1 Datasets

Three independent corpora were used for normative training, primary evaluation, and cross-disorder generalization. The VOC-ALS database (Bionetworks, 2026) comprises 153 subjects, including 102 individuals with ALS and 51 healthy controls, recorded across eight structured speech tasks consisting of five sustained vowels (/a/, /e/, /i/, /o/, /u/) and three diadochokinetic syllables (/pa/, /ta/, /ka/), yielding 1,294 recordings. The corpus has previously supported computational analysis of bulbar dysfunction in ALS (Tena et al., 2023). All audio was downsampled from 44.1 kHz to 16 kHz, consistent with established speech biomarker pipelines (Eyben et al., 2016). Normative training was restricted exclusively to the 51 healthy controls, producing 354 healthy clips; no ALS recordings were used during generative model training. Subject-disjoint five-fold stratified cross-validation with a fixed random seed of 42 was implemented, assigning subjects rather than clips to folds, and identical splits were applied across all baseline comparators to ensure fairness. For cross-disorder evaluation, the trained framework was applied to DementiaNet (Gite, 2022), containing 145 subjects and 455 spontaneous speech recordings, and the Italian Parkinson’s Voice and Speech database (Dimauro, 2019), containing 65 speakers and 831 recordings across sustained vowels, text reading, and diadochokinetic tasks. Neither external cohort contributed to model training, normalization statistics, or classifier calibration.

### 3.2 Feature Extraction

Each 16 kHz recording was converted to a log-mel spectrogram using a 400-sample Hann window, 160-sample hop length corresponding to 10 ms, a 512-point fast Fourier transform, and 128 mel filter banks spanning 0–8 kHz. After truncation or zero-padding to 256 frames, each clip was represented as a 128 × 256 time–frequency matrix. Log-mel representations are widely adopted in pathological speech modeling because they preserve perceptually relevant spectral structure while reducing dimensionality (Eyben et al., 2016; Song et al., 2024). Mel scaling concentrates spectral resolution below 4 kHz, the band containing fundamental frequency and primary formant structure associated with neuromotor control of the laryngeal and articulatory systems, including the nucleus ambiguus and hypoglossal nucleus (Yunusova et al., 2012). To prevent information leakage, zero-mean unit-variance normalization was performed using statistics computed exclusively from healthy training clips within each fold.

### 3.3 Feature Encoding and Conditioning

Each spectrogram was paired with a two-dimensional conditioning vector consisting of age and sex. Age was standardized using the mean and standard deviation derived solely from healthy training subjects within each fold, preventing test-fold information from influencing the normative reference. Sex was encoded as 0 for male and 1 for female, with missing values set to 0.5. For DementiaNet, age metadata was unavailable and therefore fixed at 0.5, while sex was encoded where available. Demographic conditioning was included because baseline vocal fold vibration, formant dispersion, and perturbation measures vary systematically with age and sex (Kent et al., 1999; Wuyts et al., 2000), and failure to account for these effects can confound physiological variation with pathological deviation.

### 3.4 Normative Model Architecture

SPEAK-NORM employs a conditional variational autoencoder to learn an age- and sex-adjusted distribution of healthy motor speech, as illustrated in Figure 1. The architecture consists of a convolutional encoder that compresses each 128 × 256 spectrogram into a 32-dimensional latent representation and a mirrored convolutional decoder that reconstructs the input. Normative modeling approaches have demonstrated value in separating disease-specific deviation from healthy population variability in computational medicine (Bozek et al., 2023), and conditional generative models such as cVAEs are well suited for capturing structured biological variation (Ram-chandran et al., 2023). The model was trained exclusively on healthy control clips within each fold for 25 epochs using the Adam optimizer with a learning rate of 0.001 and batch size of 64, with early stopping based on validation reconstruction loss. No ALS, dementia, or Parkinson’s disease recordings were used to shape the learned distribution. Since the model learns the conditioned distribution of healthy motor speech rather than directly separating labeled disease groups, it evaluates each recording as a structured departure from age- and sex-adjusted normative coordination. This framework enables quantification of mild bulbar motor abnormalities through multi-domain deviation features, supporting detection of early-stage motor-speech impairment.

**Figure 1.**
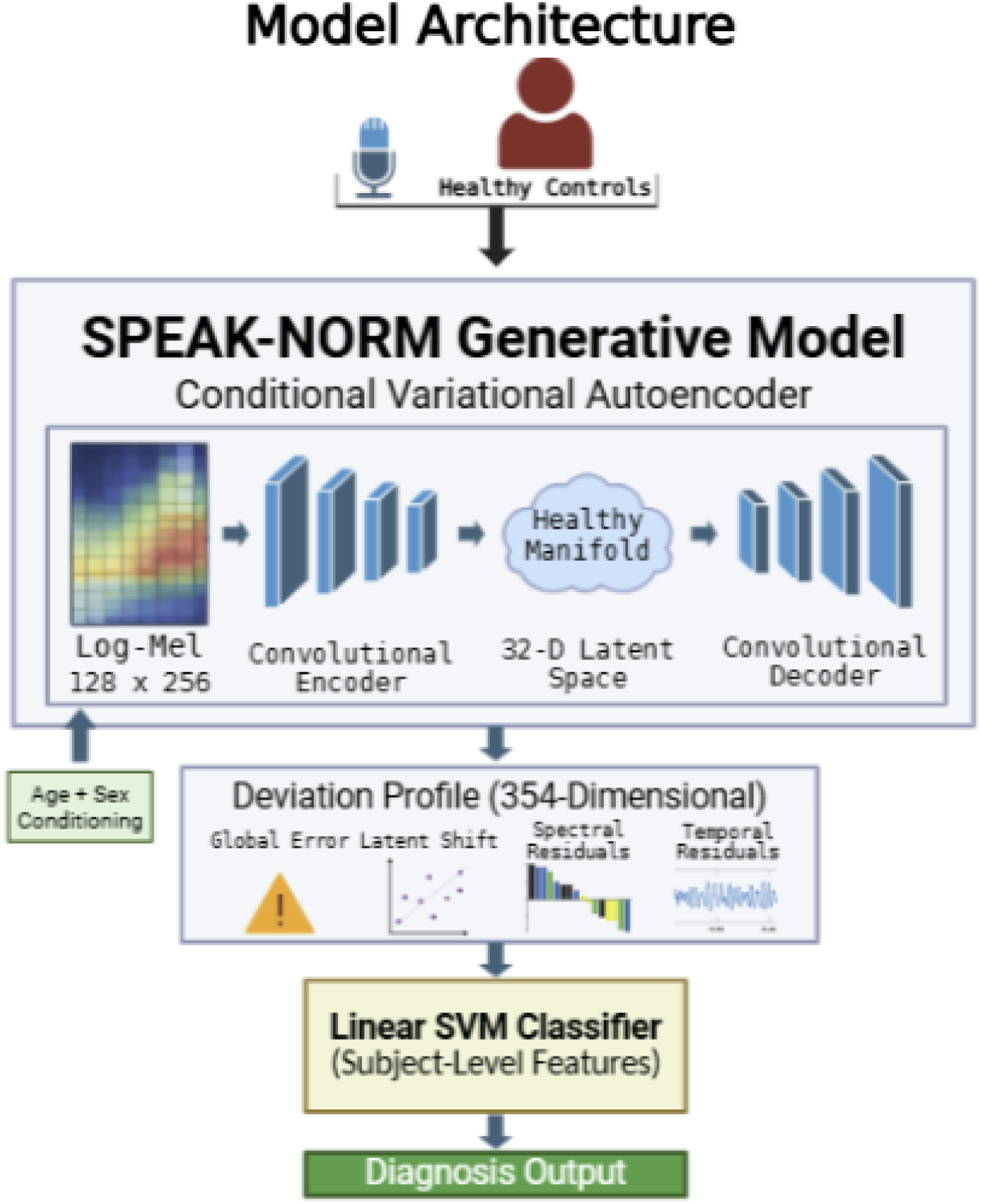
Overview of the SPEAK-NORM framework. Healthy control recordings are used to train an age- and sex-conditioned generative model of normative motor speech.

### 3.5 User-Level Deployment Workflow

As illustrated in Figure 2, SPEAK-NORM is designed for deployment using standard consumer recording devices. A speech sample can be collected using a smartphone or laptop microphone without the need for specialized clinical instrumentation such as electromyography systems or proprietary acoustic analysis platforms. The recording is processed to generate interpretable deviation summaries that quantify structured departures across global, latent, spectral, and temporal acoustic domains, alongside calibrated confidence scores that express diagnostic certainty. By eliminating reliance on hardware-dependent clinical voice analysis tools, the framework enables low-cost, scalable neuromotor screening in both clinical and resource-limited environments.

**Figure 2.**
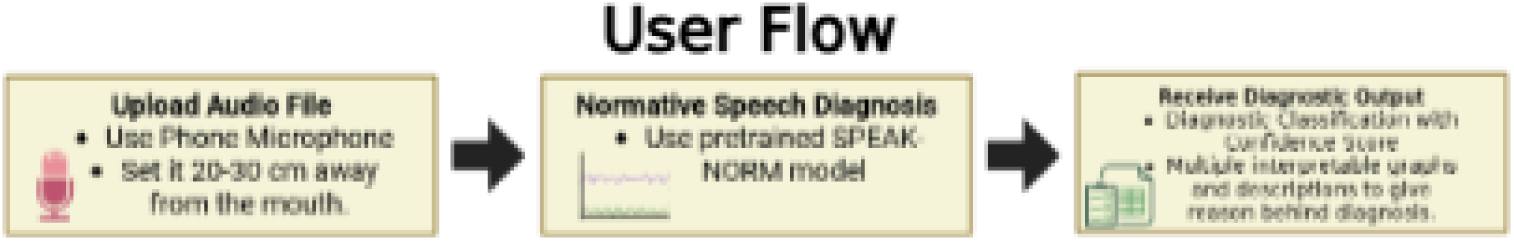
User-level deployment workflow for SPEAK-NORM. A speech recording is processed through normative evaluation to produce subject-level di-agnostic probability.

### 3.6 Deviation Scoring and Classification

Following normative training, each recording was transformed into a 354-dimensional deviation profile quantifying structured departure from the healthy reference across global, latent, temporal, and spectral dimensions. The representation integrates overall reconstruction error, a 32-dimensional latent mean vector, latent-space distance from the healthy distribution, reconstruction error aggregated across time, reconstruction error aggregated across frequency, and structured residual patterns capturing localized deviations. This multi-domain deviation strategy aligns with prior work demonstrating that structured acoustic characterization improves discrimination of dysarthria and neurodegenerative speech phenotypes (Liss et al., 2010; Tena et al., 2023). All deviation components were standardized using healthy training statistics within each fold. Subject-level representations were obtained by averaging clip-level deviation profiles across tasks, and a linear support vector classifier with class-balanced weighting was trained on these subject-level features, consistent with established ALS speech classification pipelines (Tena et al., 2023). Probability calibration was performed using internal cross-validation restricted to training data.

### 3.7 Statistical Analysis

Performance was evaluated using subject-disjoint five-fold stratified cross-validation, with ROC-AUC, PR-AUC, accuracy, F1-score, balanced accuracy, sensitivity, specificity, and precision reported in accordance with established standards in pathological speech modeling (Agurto et al., 2024). Statistical significance was assessed using permutation testing by randomly shuffling training labels within each fold and re-executing the complete pipeline, thereby evaluating the null hypothesis of no association between deviation features and diagnostic labels (Bozek et al., 2023). To determine whether the learned representations captured disorder-specific structure rather than generic neurodegeneration, healthy-normalized deviation vectors were computed separately for ALS, dementia, and Parkinson’s disease cohorts using dataset-specific normative models. Feature-wise effect sizes and block-level summaries reflecting reconstruction error, latent shifts, spectral deviation, and temporal deviation were compared across disorders using nonparametric statistical testing. Within the ALS cohort, stratification capacity was examined by applying unsupervised clustering to subject-level deviation vectors to identify motor-speech phenotype groups. For interpretability, subjects were ordered by cluster assignment and secondarily by ALSFRS-R speech subscore and mean absolute deviation, and visualized using a two-panel heatmap in which compact deviation summaries were displayed alongside aligned ALSFRS-R speech subscores. When predefined summary blocks (GlobalRecon, LatentShift, SpectralResidual, TemporalResidual, DeviationMagnitude) were available they were used directly; otherwise, robust aggregate summaries were computed from the deviation feature columns. Deviation summaries were z-scored across ALS subjects for visualization only.

## 4 Results and Discussion

### 4.1 Data

#### 4.1.1 ALS Diagnosis Performance

SPEAK-NORM achieves the highest classification performance compared to other State-of-The-Art baselines under subject-disjoint evaluation, as summarized in Table 1. The model uses 354-dimensional deviation features extracted from the normative cVAE and a class-balanced linear SVM with probability calibration. During five-fold stratified cross-validation on the training split, subjects were exclusively assigned to either training or validation folds to prevent identity leakage. Clip-level deviation profiles were averaged to produce subject-level representations prior to classification, reflecting clinical decision-making at the patient level.

**Table 1.** Subject-level ALS diagnosis performance under identical subject-disjoint five-fold cross-validation.

| Metric | Tena et al. | Ilias et al. | Agurto et al. | SPEAK-NORM |
| --- | --- | --- | --- | --- |
| Accuracy (%) | 73.1 | 82.7 | 76.9 | **98.1** |
| F1-score (%) | 81.3 | 81.4 | 78.1 | **98.6** |
| Sensitivity (%) | 79.0 | 80.2 | 81.3 | **98.5** |
| Specificity (%) | 60.0 | 84.7 | 72.5 | **97.1** |

To evaluate robustness, a separate held-out test split was constructed, and the final calibrated SVM was trained on all training subjects before evaluation on unseen test subjects. Performance on the hold-out set closely matched cross-validation results, indicating minimal overfitting and stable generalization. The difference between mean cross-validation ROC-AUC and held-out ROC-AUC was small, confirming consistency between internal validation and external testing.

As illustrated in Figure 3, misclassification rates remain low on the held-out subjects. Healthy controls are rarely predicted as ALS, and ALS subjects are rarely predicted as healthy, demonstrating balanced screening performance rather than asymmetric class bias. The confusion matrix reflects true subject-level predictions after aggregation across clips.

**Figure 3.**
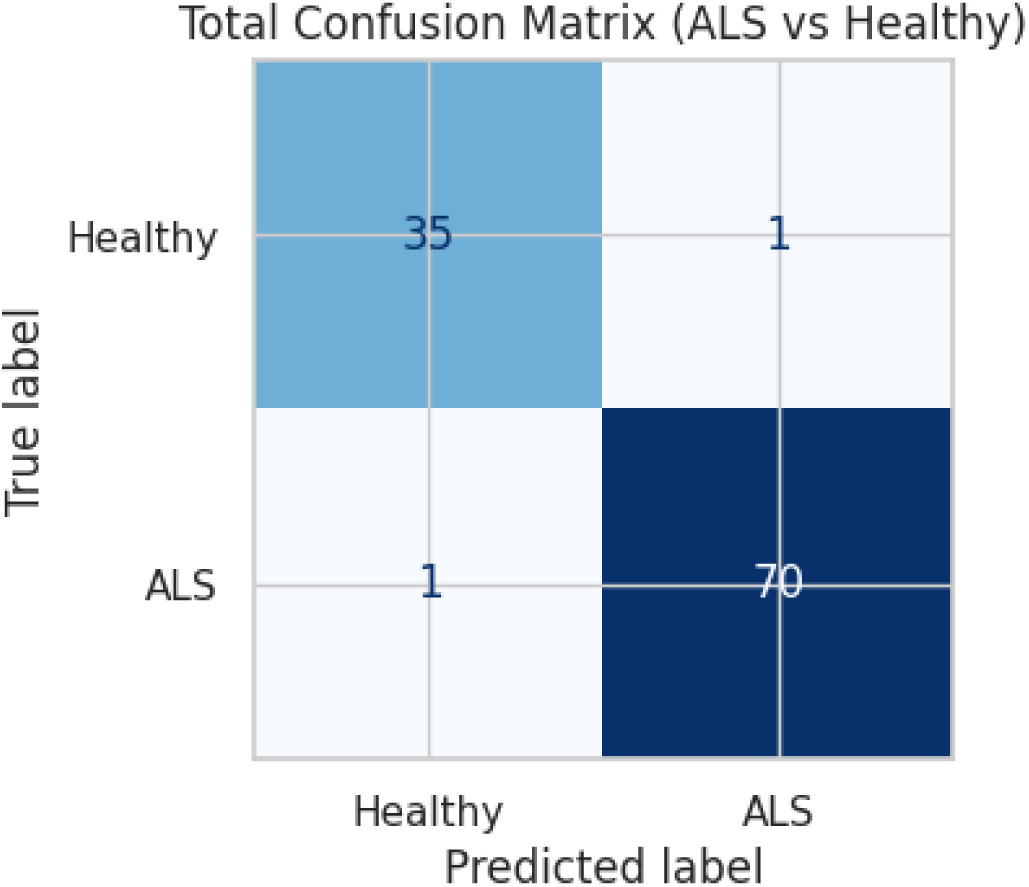
Held-out subject-level confusion matrix for ALS versus healthy classification using SPEAK-NORM.

Together, these results demonstrate that deviation from an age- and sex-conditioned healthy speech manifold provides stable and reproducible discrimination of ALS under clinically realistic subject-level evaluation.

#### 4.1.2 Comparison with Existing Clinical Methods

The performance of SPEAK-NORM was compared against established clinical acoustic indices commonly used in ALS and dysarthria assessment, including eGeMAPS (Eyben et al., 2016), Dysphonia Severity Index (DSI) (Wuyts et al., 2000), F2 slope analysis (Yunusova et al., 2012), Envelope Modulation Spectrum (EMS) features (Liss et al., 2010), and Formant Centralization Ratio with Bark-scaled Vowel Space Area (FCR + VSA-Bark) (Sapir et al., 2010). Each method was evaluated under identical subject-disjoint conditions to ensure fair comparison.

The eGeMAPS feature set extracts 88 predefined acoustic descriptors spanning spectral energy distribution, cepstral coefficients, formant characteristics, and perturbation measures. Features include jitter, shimmer, harmonics-to-noise ratio, MFCC statistics, and spectral slope parameters computed over entire recordings. DSI combines four scalar measures, jitter, shimmer, maximum phonation time, and highest fundamental frequency, into a weighted composite severity score originally designed for sustained vowel assessment. F2 slope quantifies the rate of second formant transition during articulatory movements, reflecting lingual motor velocity during vowel changes. EMS analyzes amplitude modulation spectra to capture rhythmic and temporal irregularities in diadochokinetic tasks. FCR + VSA-Bark compute geometric relationships among vowel formants to estimate articulatory centralization, measuring reductions in vowel space associated with dysarthria.

All baseline approaches rely on handcrafted scalar descriptors extracted independently from either sustained phonation or repetitive syllable tasks. These metrics summarize isolated acoustic properties rather than modeling coordinated multi-pathway motor activity across spectral, temporal, and latent domains.

As summarized in Table 2, balanced accuracy for these methods ranges from 0.505 to 0.627 under subject-disjoint evaluation. In contrast, SPEAK-NORM achieves a balanced accuracy of 0.978, with sensitivity of 0.985 and specificity of 0.971. The improvement exceeds 35 percentage points in balanced discrimination compared to all clinical baselines. Importantly, gains are observed simultaneously in both sensitivity and specificity, indicating that improvement is not driven by threshold adjustment but by increased separation between ALS and healthy subjects in the learned deviation space.

**Table 2.** Comparison of SPEAK-NORM with established clinical acoustic in-dices under identical subject-disjoint evaluation.

| Method | bAcc | Sens | Spec | Prec |
| --- | --- | --- | --- | --- |
| eGeMAPS | 0.627 | 0.490 | 0.764 | 0.841 |
| DSI | 0.532 | 0.364 | 0.700 | 0.612 |
| F2 Slope | 0.505 | 0.668 | 0.342 | 0.685 |
| EMS | 0.542 | 0.492 | 0.591 | 0.692 |
| FCR + VSA-Bark | 0.509 | 0.387 | 0.631 | 0.525 |
| **SPEAK-NORM** | **0.978** | **0.985** | **0.971** | **0.988** |

Figure 4 illustrates that performance gaps persist across accuracy, ROC-AUC, and F1-score, demonstrating that improvements are consistent across threshold-independent and threshold-dependent metrics. These findings indicate that modeling structured deviation from an age- and sex-conditioned healthy distribution provides substantially stronger ALS discrimination than scalar perturbation metrics derived from isolated acoustic parameters.

**Figure 4.**
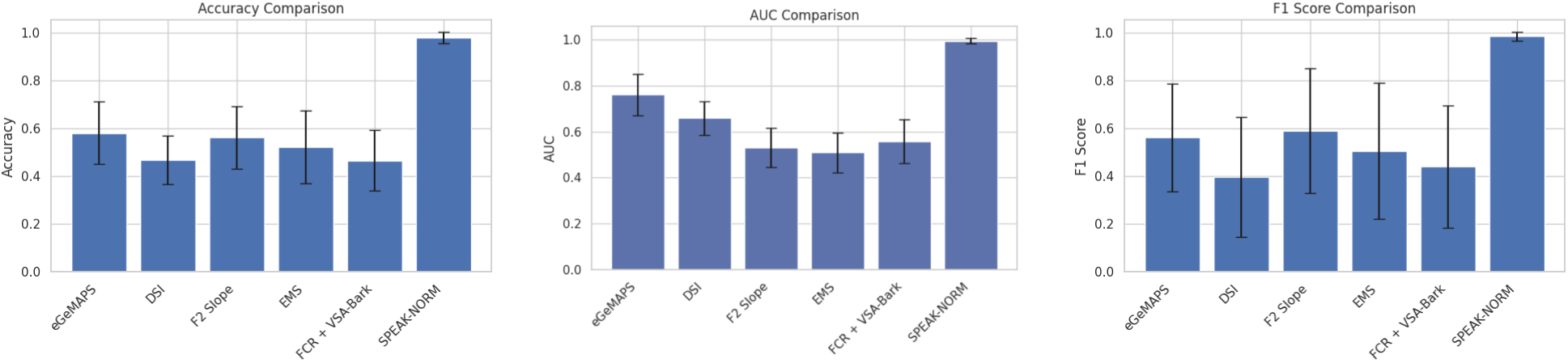
Comparison of SPEAK-NORM with established clinical acoustic indices across accuracy, ROC-AUC, and F1-score under subject-disjoint evaluation.

#### 4.1.3 Disease-Specific Deviation Analysis

To determine whether SPEAK-NORM captures ALS-specific motor pathway degeneration rather than generic neurodegenerative impairment, cross-disorder evaluation was conducted on independent dementia and Parkinson’s disease cohorts. For each external dataset, the conditional generative model was retrained exclusively on that dataset’s healthy controls to eliminate recording-condition and language variability, after which deviation features were computed and classified without exposure to pathological samples from other disorders.

As shown in Table 3, the framework maintains high accuracy across disorders, indicating that deviation modeling generalizes beyond ALS while remaining sensitive to disease-specific acoustic structure. Importantly, cross-disorder evaluation does not involve transferring ALS-trained classifiers; instead, each dataset is evaluated relative to its own healthy reference distribution. This design isolates pathological deviation from demographic and recording confounds.

**Table 3.** Classification accuracy of SPEAK-NORM across neurodegenerative cohorts using dataset-specific normative retraining.

| Dataset | Accuracy (%) |
| --- | --- |
| ALS (VOC-ALS) | **98.14** |
| DementiaNet | 89.1 |
| Parkinson’s Voice Dataset | 98.75 |

Feature-level analysis reveals that deviation patterns differ systematically across disorders. Figure 5 presents effect size distributions across global reconstruction error, latent shift magnitude, spectral residuals, and temporal residuals. ALS demonstrates dominant temporal and latent deviations, consistent with degeneration of hypoglossal and laryngeal motor neurons affecting articulatory timing and coordination. Parkinson’s disease shows stronger spectral residual shifts, reflecting hypokinetic phonation and reduced amplitude modulation. Dementia exhibits moderate, distributed deviation without the pronounced temporal instability observed in ALS. Nonparametric comparisons across blocks yield statistically significant differences (*p <* 0.05), confirming structured dissociation rather than uniform degradation.

**Figure 5.**
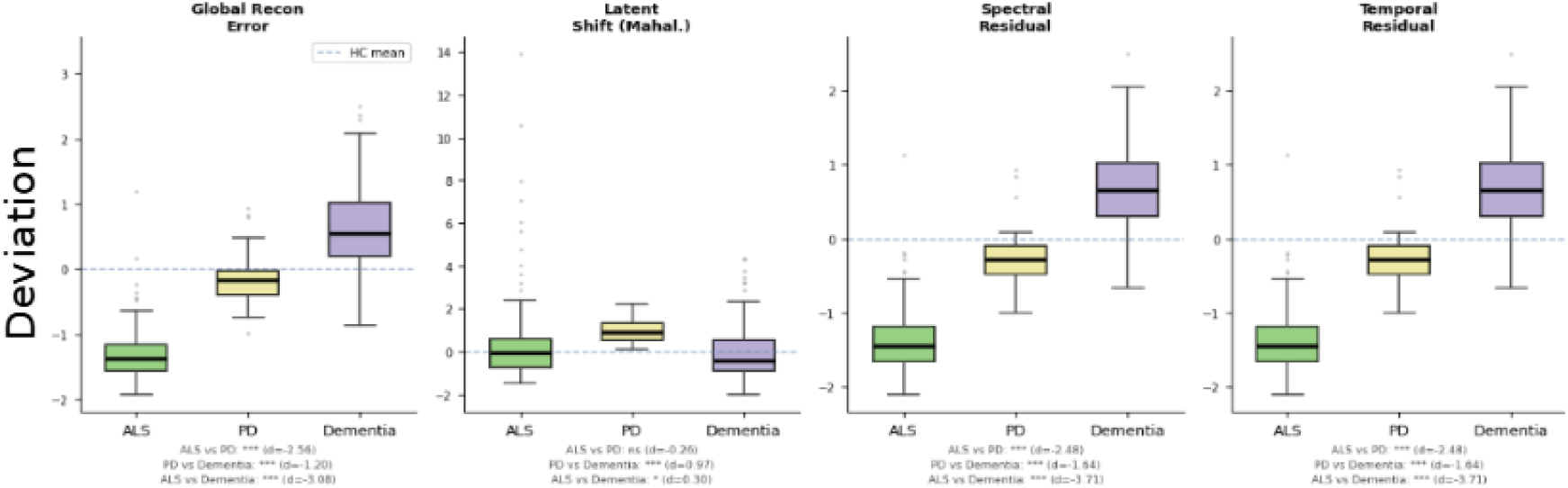
Block-level deviation comparison across ALS, dementia, and Parkinson’s disease. Distinct effect size patterns indicate disorder-specific acoustic deviation profiles.

To further quantify cross-disorder separability, the similarity of top-ranked deviation features was assessed using Jaccard similarity. As shown in Figure 6, ALS shares minimal overlap with either dementia or Parkinson’s disease (off-diagonal similarity *≤* 0.143), indicating that the highest-contributing deviation dimensions are largely disorder-specific. This low overlap demonstrates that the model does not rely on generic severity signals but instead activates distinct acoustic dimensions across pathologies.

**Figure 6.**
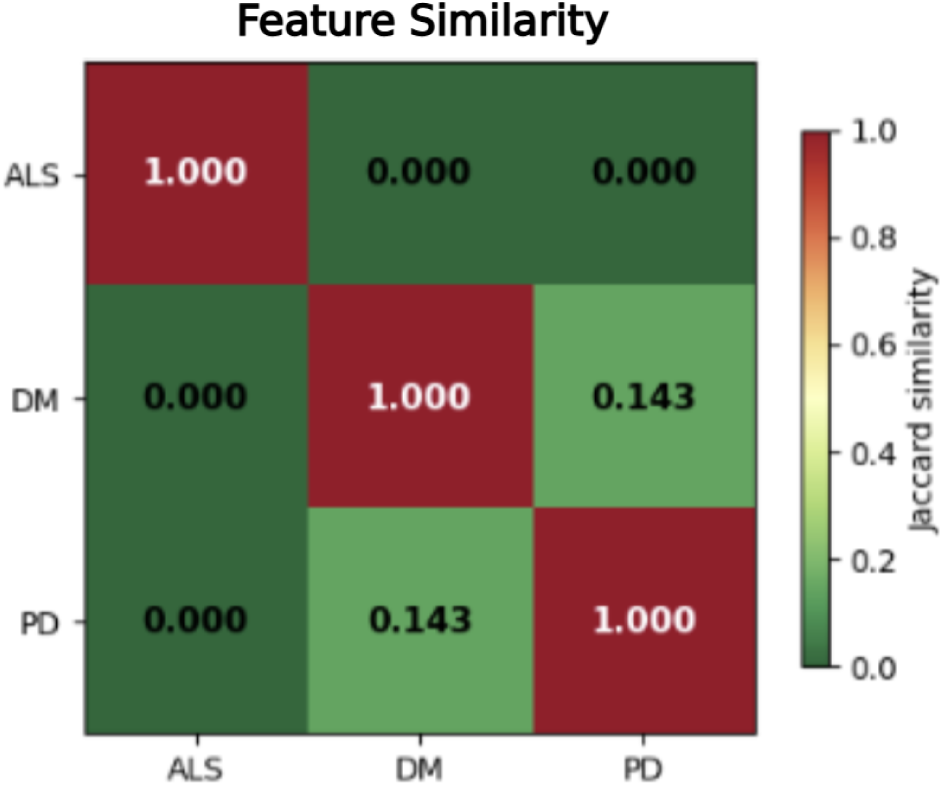
Top-20 feature ranking similarity across disorders. Low off-diagonal values indicate disorder-specific deviation dimensions.

The feature importance heatmap in Figure 7 further illustrates structured dissociation. Parkinson’s disease exhibits the strongest spectral and timing residual magnitudes, dementia shows moderate latent and spectral activation, and ALS demonstrates balanced but temporally weighted deviation. The diagonal dominance pattern confirms that each disorder activates a distinct subset of deviation blocks.

**Figure 7.**
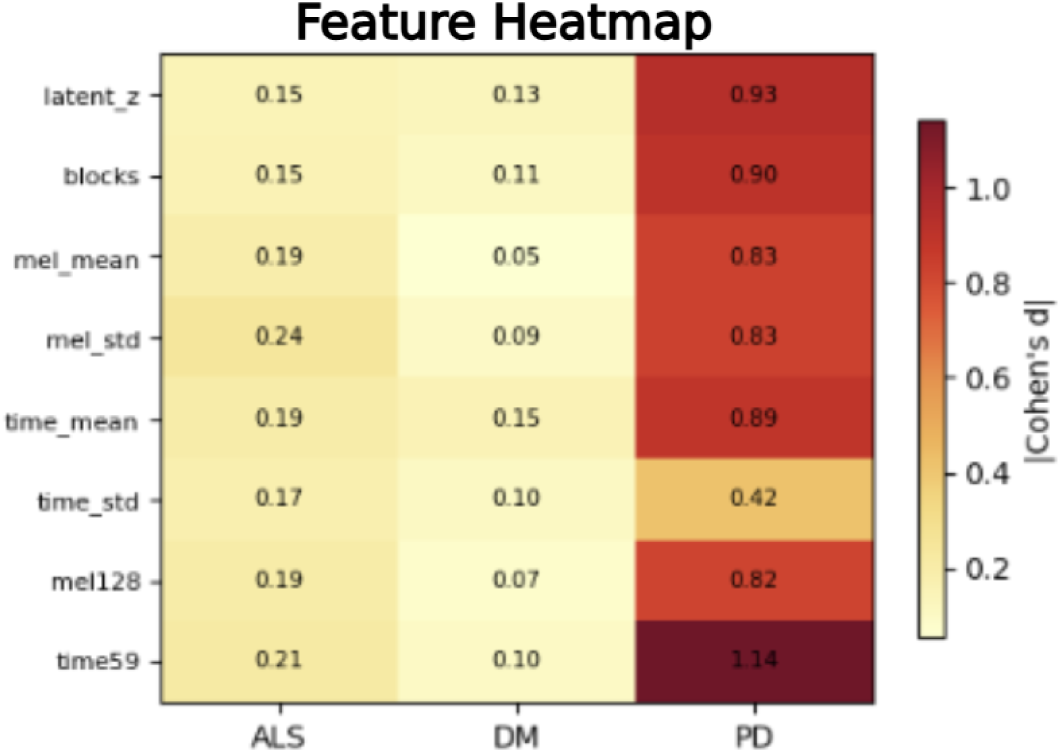
Block-level deviation heatmap (Cohen’s d) across disorders. Distinct activation patterns support disease-specific acoustic signatures.

Together, these results demonstrate that SPEAK-NORM does not learn a generic “neurodegeneration” signal. Instead, it models structured departure from demographic-conditioned healthy motor speech and reveals pathway-specific acoustic signatures that differ across ALS, Parkinson’s disease, and dementia.

#### 4.1.4 Alignment with Clinically-Accepted Scores

To evaluate clinical relevance, subject-level deviation profiles were compared with ALSFRS-R speech subscores within the ALS cohort. As shown in Figure 8, subjects with lower ALSFRS-R speech scores, indicating greater bulbar impairment, exhibit consistently elevated deviation across global reconstruction error, latent shift magnitude, and residual dimensions. In contrast, subjects with preserved speech function display lower and more uniform deviation patterns. The ordered heatmap demonstrates a clear gradient, with deviation magnitude increasing alongside clinical severity rather than varying randomly across individuals. This structured alignment indicates that SPEAK-NORM deviation features reflect clinically recognized motor-speech impairment and capture progressive bulbar involvement consistent with ALSFRS-R assessment.

**Figure 8.**
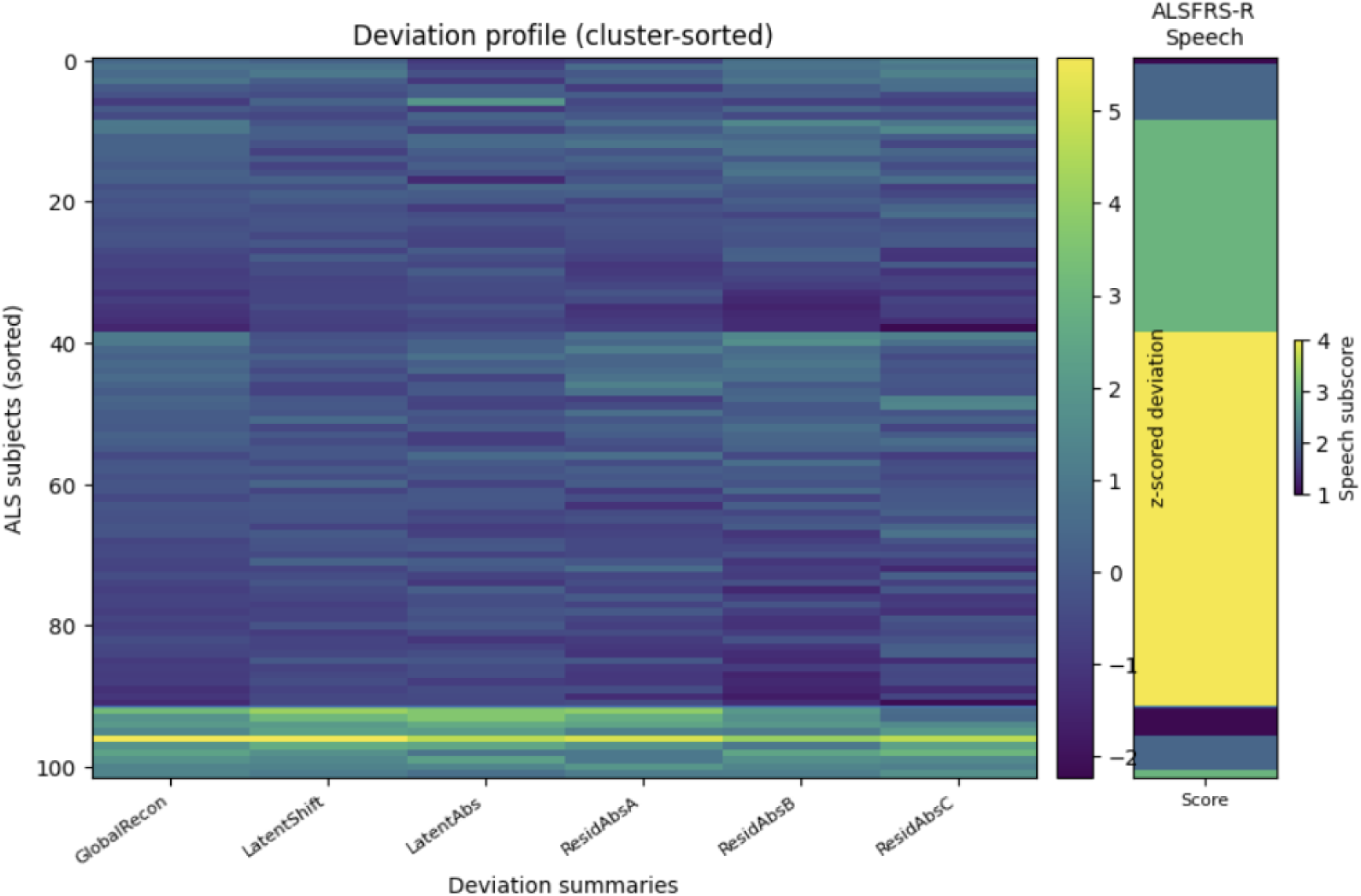
Subject-level deviation profiles (left) aligned with ALSFRS-R speech subscores (right). Subjects are cluster-sorted to highlight structured variation across motor-speech impairment.

#### 4.1.5 Early Diagnosis Potential

To assess sensitivity to early-stage impairment, binary classification experiments were conducted across severity strata derived from ALSFRS-R speech scores. As shown in Figure 9, discrimination between healthy and moderate impairment reached 95% accuracy, while mild versus moderate classification achieved 89% accuracy. Importantly, separation between healthy and mild cases achieved 83% accuracy, demonstrating that the deviation representation detects impairment prior to severe bulbar decline. These results indicate that structured deviation features capture early motor-speech disruption rather than only advanced dysarthria. The performance gap between healthy–moderate and healthy–mild comparisons reflects the expected clinical continuum, where early-stage abnormalities are subtler but still measurable. Unlike threshold-based perturbation metrics, which typically detect only established dysarthria, normative deviation modeling identifies structured deviations consistent with emerging bulbar involvement. These findings support the potential of SPEAK-NORM as an early screening tool for progressive motor neuron degeneration.

**Figure 9.**
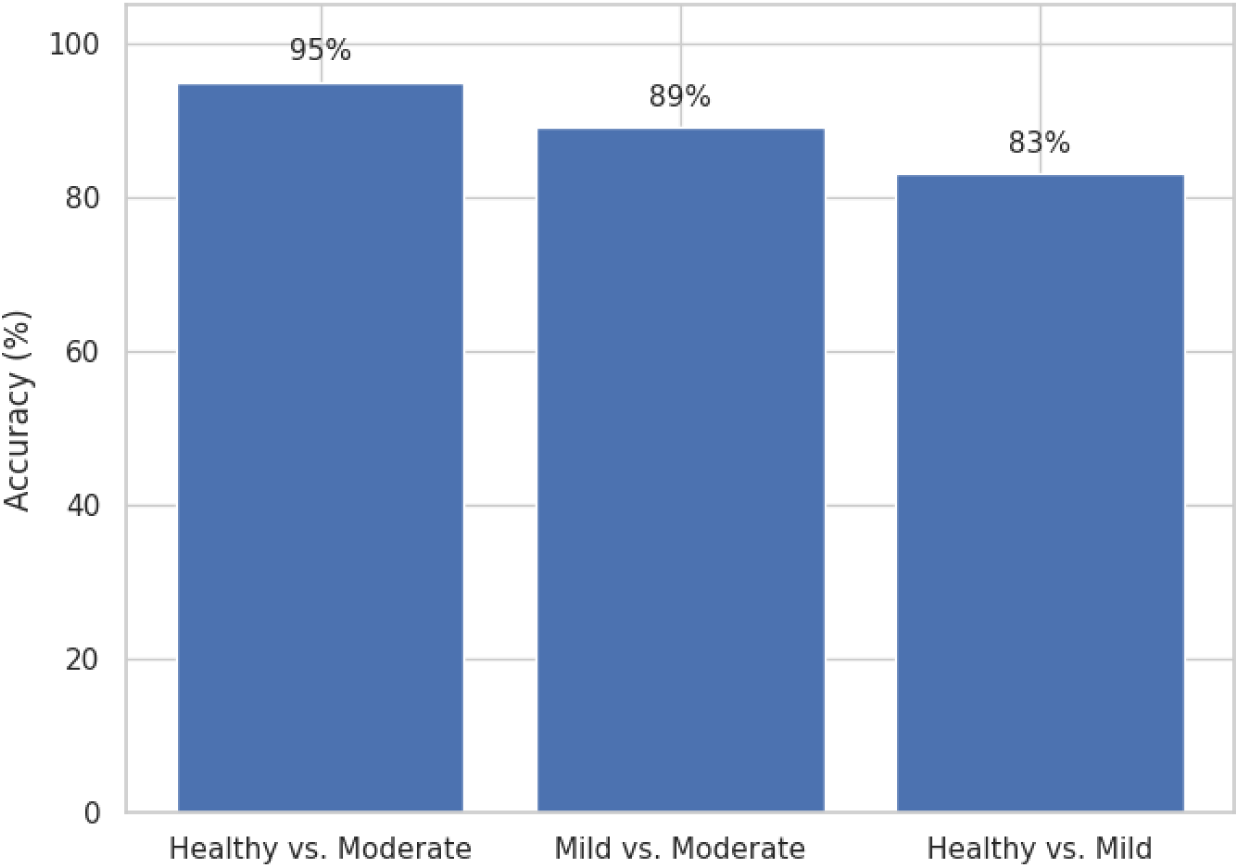
Binary severity-stratified classification performance demonstrating early detection capability.

### 4.2 Data Analysis and Discussion

Bulbar motor neuron degeneration in ALS manifests as progressive alterations in articulatory timing, phonatory stability, and respiratory support.

Figure 10 provides a detailed example of the normative modeling mechanism, showing the original log-mel spectrogram, the generative reconstruction, and the squared residual error map highlighting localized deviations. In healthy speech, spectral energy transitions are smooth and temporally coordinated. In mild impairment, localized instability emerges, particularly in high-frequency bands and temporal envelope fluctuations. In severe impairment, sustained spectral distortion and temporal irregularity become pronounced, reflecting reduced neuromuscular coordination and impaired motor unit recruitment.

**Figure 10.**
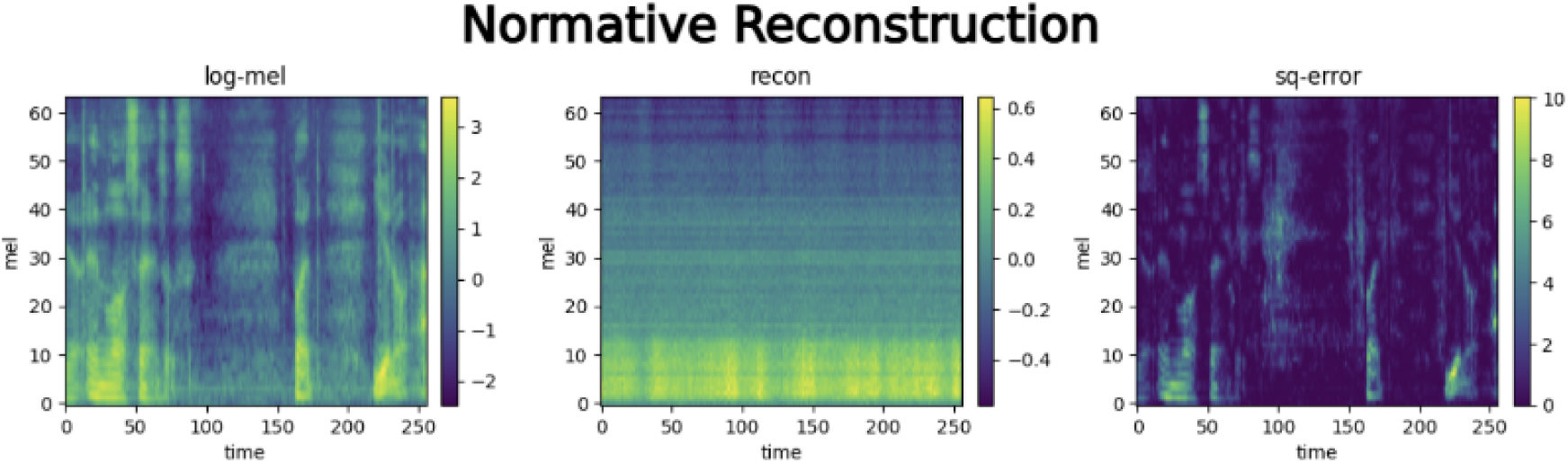
Normative reconstruction and deviation localization in a representative speech sample. **Left:** Log-mel spectrogram of the observed recording. **Middle:** Age- and sex-conditioned reconstruction generated by the cVAE trained on healthy speech. **Right:** Squared reconstruction error map highlighting localized spectral and temporal deviations from the normative manifold. Regions of elevated residual energy indicate structured departure from healthy motor-speech coordination.

The SPEAK-NORM framework models this progression by learning an age- and sex-conditioned distribution of healthy motor speech and quantifying structured departure from it. Unlike threshold-based acoustic metrics that rely on scalar perturbation measures such as jitter or shimmer, the proposed approach captures multidimensional deviation across latent, spectral, and temporal domains simultaneously. This allows the model to identify coordinated disruption across bulbar motor pathways rather than isolated perturbation events. The progressive amplification of deviation magnitude shown in Figure 11 aligns with known degeneration of the hypoglossal nucleus, nucleus ambiguus, and respiratory motor neurons, which collectively influence articulatory precision, vocal fold stability, and breath support.

**Figure 11.**
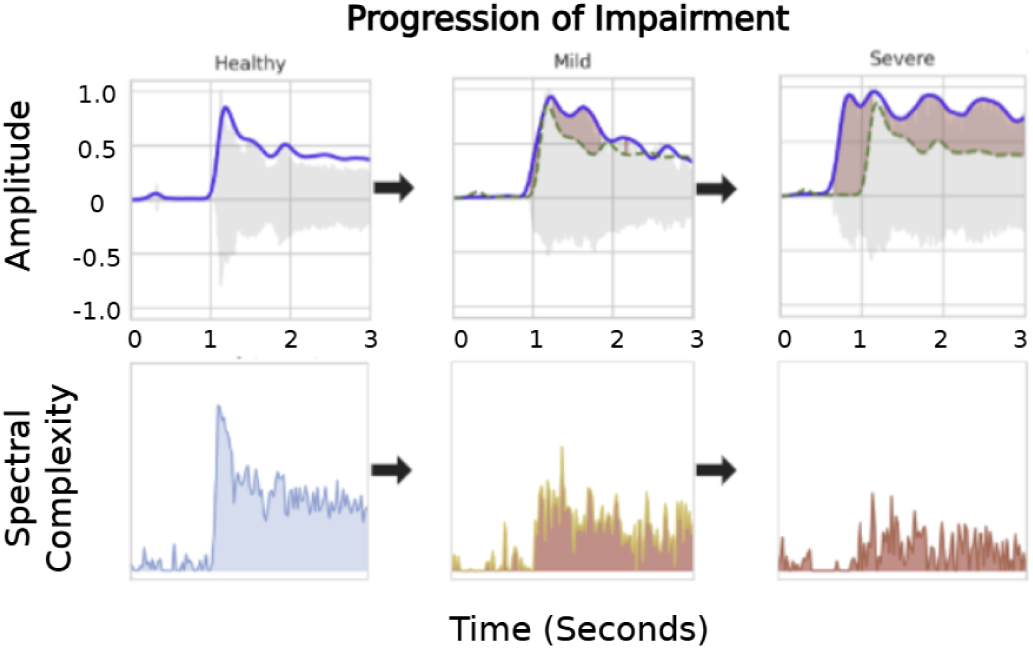
Progressive motor-speech impairment illustrated through normative reconstruction and deviation profiles. Each column represents (A) Healthy, (B) Mild, and (C) Severe conditions. Top row: Observed spectrogram (solid blue) overlaid with normative reconstruction (dashed green). Healthy speech shows close alignment, while mild impairment introduces localized spectral and temporal mismatch. Severe impairment exhibits sustained distortion and broad deviation from the healthy reference. Bottom row: Corresponding deviation magnitude profiles, where warmer regions indicate greater departure from normative motor coordination. The progressive increase in structured residuals reflects worsening bulbar motor dysfunction.

The model demonstrates strong performance in separating healthy subjects from individuals with ALS. From a clinical perspective, minimizing false negatives is critical in a progressive and life-limiting disease. The subject-disjoint evaluation indicates that misclassification of ALS as healthy is rare, supporting the potential utility of normative deviation modeling as a screening adjunct. At the same time, the framework preserves balanced specificity, reducing the likelihood of false-positive referral due to benign vocal variability.

### 4.3 Error Analysis

Challenges remain in distinguishing very early or mild impairment from healthy speech. As observed in severity-stratified analyses, the boundary between healthy and mild bulbar dysfunction is less distinct than that between healthy and moderate impairment. This mirrors clinical reality, where early dysarthria may present subtly and variably across patients. Differences in symptom onset patterns, such as whether degeneration initially affects lingual articulation versus phonatory control, may produce heterogeneous acoustic signatures that complicate early categorization.

Continuous quantification of deviation magnitude offers potential advantages over discrete clinical scoring systems. While ALSFRS-R provides ordinal categories of speech function, deviation modeling yields a continuous representation of motor-speech disruption. Such quantitative monitoring may enable finer-grained tracking of progression and therapeutic response. Longitudinal validation would be required to determine sensitivity to within-subject change over time.

Future work will focus on extending normative deviation modeling to true differential diagnosis within a single unified dataset containing ALS, Parkinson’s disease, frontotemporal dementia, and healthy controls recorded under identical protocols. Although cross-disorder experiments in this study retrained normative references using healthy subjects within each external dataset to eliminate recording and language variability, a shared multi-disease corpus would allow direct head-to-head classification and clearer assessment of disorder-specific deviation structure under controlled acquisition conditions. Such a dataset would enable evaluation of whether disease-specific latent dimensions remain separable when acoustic variability from hardware, language, and recording environment is minimized.

An additional priority is cross-linguistic generalization. Motor pathway degeneration affects articulatory coordination, phonatory stability, and respiratory timing in ways that are not language-specific, but phoneme inventories and prosodic structure vary substantially across languages. Training language-conditioned normative models or multilingual generative references would allow testing whether deviation signatures transfer across English, Spanish, Italian, and other linguistic contexts. Demonstrating consistent deviation structure across languages would strengthen the biological validity of the framework and expand its applicability to global populations.

Despite promising results, the present study relies on publicly available speech corpora and retrospective evaluation. Expansion to larger, prospectively collected, multi-center datasets will be necessary to establish robustness in real-world screening environments. Future investigations may also integrate longitudinal recordings, multimodal respiratory or articulatory sensors, and adaptive generative architectures to further refine early detection and phenotype stratification of bulbar involvement in ALS.

## 5 Conclusion

This study introduces a normative speech modeling framework for ALS diagnosis that quantifies structured deviation from age- and sex-conditioned healthy motor coordination. The SPEAK-NORM model achieves 98% classification accuracy under subject-disjoint evaluation, outperforming established clinical acoustic indices and prior supervised speech classifiers. By learning the distribution of healthy speech rather than memorizing disease examples, the framework captures multidimensional disruption across spectral, temporal, and latent domains. Disease-specific analyses demonstrate distinct deviation patterns across ALS, dementia, and Parkinson’s disease, while alignment with ALSFRS-R speech scores confirms clinical relevance. Severity-stratified experiments further indicate sensitivity to early-stage impairment, supporting potential use in pre-threshold detection. The reconstruction visualizations illustrate progressive amplification of deviation magnitude from mild to severe impairment, consistent with known bulbar motor neuron degeneration. The approach provides a scalable, low-cost screening tool requiring only standard recording devices, offering an accessible and objective complement to clinical assessment and longitudinal monitoring of neuromotor disease.

## Data Availability

All data used in the study is publicly available, and all links to the datasets can be found in the works cited.

https://github.com/shreyasgite/dementianet

https://ieee-dataport.org/open-access/italian-parkinsons-voice-and-speech

https://www.synapse.org/Synapse:syn53009474/wiki/624730

## Notes

### Competing Interest Statement

The authors have declared no competing interest.

### Author Declarations

The study only used publicly available human data. This data can be taken from 3 public datasets: VOC-ALS (https://www.synapse.org/Synapse:syn53009474), DementiaNet (http://github.com/shreyasgite/dementianet), and Italian Parkinson's Voice and Speech Database (https://ieee-dataport.org/open-access/italian-parkinsons-voice-and-speech).

